# Polygenic risk score analysis identifies deleterious protein-coding variants in novel immune pathway genes *ATP8B4, FCGR1A*, and *LILRB1* that associate with Alzheimer’s disease

**DOI:** 10.1101/2022.07.12.22277557

**Authors:** Joseph S. Reddy, Xue Wang, Mariet Allen, Minerva M. Carrasquillo, Joanna M. Biernacka, Gregory D. Jenkins, Brandon J. Coombes, Olivia Belbin, Todd E. Golde, Nilüfer Ertekin-Taner, Steven G. Younkin

**Affiliations:** Department of Quantitative Health Sciences, Mayo Clinic, Jacksonville, FL, 32224, USA; Department of Neuroscience, Mayo Clinic, Jacksonville, FL, 32224, USA; Department of Quantitative Health Sciences, Mayo Clinic, Rochester, MN, 55905, USA; Memory Unit and Biomedical Research Institute, IIB Sant Pau, c/Sant Quintí 77, 08041, Barcelona, Spain; Centre of Biomedical Investigation Network for Neurodegenerative Diseases (CIBERNED), Spain; Departments of Neuroscience and Neurology, McKnight Brain Institute, Center for Translational Research in Neurodegenerative Disease, University of Florida, Gainesville, Fl 32610 (and as of 08/01/2022: Departments of Pharmacology and Chemical Biology and Neurology, Center for Neurodegenerative Disease, Goizueta Alzheimer’s Disease Research Center, Emory University School of Medicine, Atlanta, GA, USA 30322; Department of Neurology, Mayo Clinic, Jacksonville, FL, 32224, USA

## Abstract

**Background:** Alterations in innate immunity are pathologically associated with and genetically implicated in Alzheimer’s disease (AD). In the whole exome sequence (WES) dataset generated by the Alzheimer’s Disease Sequencing Project (ADSP), only the previously identified p.R47H variant in the innate immunity gene, *TREM2*, shows study-wide association with risk of AD. Using a novel approach, we searched the ADSP WES data to identify additional immune pathway genes with deleterious variants that, like *TREM2*.pR47H, show strong association with AD.

**Methods:** Using polygenic risk scores (PRS) to analyze association with AD, we evaluated deleterious variants (CADD Phred-scaled score > 20) with a minor allele count of 20 or more in 228 genes comprising an immune co-expression network containing *TREM2* (CEN_*TREM2*_). A significant polygenic component composed of deleterious stop-gain and non-synonymous variants was identified, and false discovery rates were determined for the variants in this component. In genes harboring a significant variant, PRS for all variants in the genes were then analyzed.

**Results:** The PRS for the 182 deleterious variants in CEN_*TREM2*_ showed significant association with AD that was driven by 142 deleterious variants (136 non-synonymous, 6 stop-gain). In the 142 variant polygenic component, four variants had significant AD risk association: *TREM2*.pR47H, two deleterious stop-gain variants (*FCGR1A*.pR92X, and *LILRB1*.pY331X) in novel AD genes and 1 non-synonymous variant *(ATP8B4*.pG395S). Remarkably, PRS for the 36 additional variants in these four genes also showed significant association with AD. The PRS for all 40 variants in the 4 genes, showed significant, replicable association with AD and 3 additional variants in this polygenic component had significant false discovery rates: *ATP8B4*.pR1059Q, *LILRB1*.pP7P, and *LILRB1*.pY327Y.

**Conclusions:** Here, we identify 3 immune pathway genes (*ATP8B4, LILRB1*, and *FCGR1A*) with a variant that associates with AD. Like *TREM2*.pR47H, each of the variants has a minor allele frequency less than 1% and is a deleterious, protein altering variant with a strong effect that increases or decreases (*LILRB1*.pY331X) risk of AD. Additional variants in these genes also alter risk of AD. The variants identified here are ideally suited for studies aimed at understanding how the innate immune system may be modulated to alter risk of AD.

## Background

Aggregation and accumulation of the amyloid β protein (Aβ) is thought to trigger a slow insidious and complex neurodegenerative cascade [1, 2]. Yet in humans, amyloid deposition, precedes clinical dementia by many years. Indeed, cognitively normal individuals with cerebral amyloid deposition can be identified by amyloid ligand positron emission tomography [3] or by a low Aβ42/Aβ40 ratio in cerebrospinal fluid or plasma [4-6]. Thus, in principle, the long prodromal period in AD offers a window of therapeutic opportunity wherein therapy to slow or halt progression to dementia might be instituted in cognitively normal subjects with cerebral amyloid deposition. There are still gaps in our understanding about how and when the innate immune system is altered during the progression of AD, but genetic studies clearly demonstrate that altered innate immune function can modulate risk for AD. Experimental and pathological data suggest that the innate immune system may regulate amyloid deposition, toxicity of the amyloid deposits, tau pathology and neurodegeneration [7, 8].

It is well-established that heterozygous missense variants in triggering receptor expressed on myeloid cells 2 (*TREM2*) strongly increase risk of AD in populations of European [9-16] and African American ancestry [17, 18]. In the human brain, *TREM*2 is selectively expressed in microglial cells [19], and *TREM2* expression is increased both in the brains of AD patients [20-23] and in mouse models of amyloid and tau deposition [24-28]. Recent studies of mouse models indicate that *TREM2* plays an important role in regulating the response of the immune system to Aβ and tau pathologies [24, 25, 27-29]. Notably, these studies suggest that AD-linked *TREM2* variants are partial loss of function, and that this partial loss of function confers risk for AD. In contrast regulatory variants that increase *TREM2* expression may decrease AD risk [30]. We previously reported weighted gene co-expression network analysis (WGCNA) of human brains transcriptomes that identified association of a *TREM2*-containing co-expression network (CEN_*TREM2*_) with AD [31]. A WGCNA of brain expression measures from amyloid-bearing mice reported that *TREM2* is a hub gene in an AD co-expression network activated by amyloid [19]. In another study, *TYROBP*, the signaling partner of *TREM2*, was found to be a key regulator in a human immune gene regulatory network relevant to AD pathology [32]. Thus, in principle, therapies which effectively target *TREM2* and other genes in its co-expression network might halt or slow progression to dementia in cognitively normal subjects with amyloid deposition by modulating the immune response that occurs when amyloid is deposited. In an effort to identify novel AD genes co-expressed with *TREM2*, we employed a novel approach using polygenic risk scores (PRS) to explore 228 genes in a *TREM2*-containing co-expression network (CEN_*TREM2*_).

## Methods

### Study Design

This is a cross-sectional case-control study to identify novel immune genes with deleterious variants that show strong association with AD. AD patients and controls were retrospectively selected from non-Hispanic White subjects from the ADSP Umbrella Study NG00067.v7. DNA was sequenced at three large scale sequencing analysis centers; data from the Broad Institute, the largest cohort, were used as a discovery dataset (N=4,174, AD=2668, Control=1506) to construct PRS, and data from Washington University (N=3299, AD=1703, Control=1596) and/or Baylor University (N=2273, AD=1188, Control=1085) were used as test datasets. Subject demographics are shown in Additional File 1.

### WES

Whole exome sequencing (WES) data and corresponding clinical information for 20,503 subjects in the seventh release of the from ADSP Umbrella Study (NG00067.v7) were obtained from the National Institute of Aging Genetics of Alzheimer’s Disease Data Storage Service after approval. Using PLINK v2.00a3LM [33], 8,152,281 variants obtained from 10,291 non-Hispanic White subjects were extracted from project level VCF files generated by Variant Calling Pipeline and data management tool [34] developed by the Genome Center for Alzheimer’s Disease.

### Quality control

Bi-allelic variants passing GATK [35] VQSR filter, having a genotyping rate ≥ 98%, a minor allele count ≥ 20, and a Hardy-Weinberg p-value > 5×10^−8^ in controls were retained. Samples with a minimum call rate of 95% and those passing sex check with an inbreeding coefficient of the X-chromosome for males>0.7 and females <0.3 were retained. Participants with a heterozygosity rate beyond 6 standard deviations (sd) from the mean were excluded. Subsequently, samples were evaluated for relatedness using KING [36] implemented via PLINK (v2.00a3LM), and retaining only one sample from each pair or family of 1^st^, 2^nd^ or 3^rd^ degree relatives. Principal component analysis was performed on samples after resolving relatedness to evaluate population substructure and exclude population outliers using Eigenstrat [37, 38]. Eigenstrat was set to remove outliers up to 6 standard deviations (sd) for the top 10 principal components (PCs) over 6 iterations, while refitting PCs after each iteration of outlier removal. In summary, 35 samples with a call rate less than 95%, 11 with discordant sex, 43 with heterozygosity beyond 6 sd, 151 relateds and 305 population outliers were excluded. A total of 151,571 variants and 9,746 participants passed filters and QC. Variants were annotated using ANNOVAR [39]. Precomputed Combined Annotation Dependent Depletion (CADD) Phred-scaled scores (v1.6) [40, 41] were obtained from https://cadd.gs.washington.edu/download.

### Genome-Wide Association Study (GWAS)

To evaluate the association of individual genetic variants with AD in all qualifying participants (N=9,746), we performed logistic regression in PLINK (v1.9) using an additive model while adjusting for sex, *APOE* ε2 and ε4 dose, sequencing centers and the first three PCs accounting for population substructure. Since the ADSP cohort was enriched for older controls (Additional File 1), we did not include age as a covariate in the regression model as correcting for age when individuals with AD are younger than controls leads to the model incorrectly inferring the age effect on AD risk, resulting in loss of statistical power [42]. Variants in the *APOE* region [43] (chr19: 44,495,939-45,296,742; GRCh38) were excluded from this analysis.

### Polygenic Risk Score (PRS) analysis

We estimated the cumulative weighted burden for a set of variants equivalent to the strategy used for polygenic risk scores. However, instead of applying the traditional PRS strategy which estimates risk across the entire genome, we focused on deleterious variants in genes and sets of genes within CEN_*TREM2*_. To construct PRS, individual variant beta coefficients were estimated in participants sequenced at the Broad Institute (discovery; N=4,174). These beta coefficients were then utilized to construct PRS for test sets that included participants sequenced at WashU in St. Louis (N=3,299) and/or at Baylor University (N=2,273). Beta estimates for variants in the discovery cohort were calculated using logistic regression in PLINK (v1.9) while adjusting for sex, *APOE* ε2 and ε4 dose and the first three PCs. Using the clump function in PLINK (v1.9), variants were pruned to reduce linkage disequilibrium (r^2^ < 0.2). These pruned betas were then used to construct PRS in the test set using R (v4.0.3) and tested for association with AD using a logistic regression, while adjusting for sex, *APOE* ε2 and ε4 dose and the first three PCs. The predictive value of the PRS to discern AD cases from cognitively normal controls was evaluated using the area under the receiver operating characteristic curve calculated using the ‘*pROC*’ package in R v4.0.3.

### Expression data

Weighted gene co-expression network analysis (WGCNA) was performed using R package WGCNA [44] to identify co-expressed genes in an RNA sequencing (RNAseq) dataset. The cohort, generation of RNAseq data and quality control steps have been described previously [45, 46]. Briefly, RNA was isolated from temporal cortex tissue of neuropathologically diagnosed AD patients and controls. RNA libraries were generated using the TruSeq RNA Sample Prep Kit (Illumina, San Diego, CA) and sequenced on an Illumina HiSeq2000 (101bp PE) multiplexing 3 samples per lane. Raw reads were aligned to GRCh37 and were counted for each gene through the MAP-RSeq pipeline [47]. Gene read counts were normalized using conditional quantile normalization [48]. After QC, 80 AD and 76 control samples were retained for analysis. To account for covariates, expression residuals were obtained using multiple linear regression implemented in R, where gene expression was the dependent variable, and sex, age at death, flow cell and RNA integrity number were the independent variables. As previously described, co-expression analysis was performed for 13,273 TCX RNAseq transcripts (13,211 unique genes), which were expressed above background levels in both this RNAseq dataset and in an independent cohort [46]. Co-expression networks based on residuals were obtained using WGCNA function *blockwiseConsensusModules* (args: networkType=“signed”, TOMType=“signed”, power=12). For genes in each co-expression network, enriched gene ontology terms were identified by function *GOenrichmentAnalysis*. Eigengenes that represent each co-expression network were obtained from function *moduleEigengenes*. One module was identified to contain *TREM2* [31] and thus genes in this module (CEN_*TREM2*_) were selected for further study.

## Results

### Single variant analysis

When the 131,028 autosomal variants which passed quality control (QC) and had a minor allele count of twenty or more were tested for association with AD in the 9,746 participants (Additional File 1) that passed QC (see Methods), only *TREM2*.pR47H reached genome-wide significance (1.28×10^−8^).

### PRS analysis of all CEN_*TREM2*_ variants

Of the 9,746 post-QC samples, 4,174 (43%) were sequenced at the Broad Institute, 3,299 (34%) at WashU in St. Louis, and 2,273 (23%) at Baylor University (Additional File 1). To evaluate variants in CEN_*TREM2*_ for association with AD, we performed PRS analysis utilizing Broad participants for discovery and the combined WashU and Baylor set for testing, as fully described in Methods. The PRS for the 1,334 independent variants (r^2^≤0.2) located in the 228 genes of CEN_*TREM2*_ showed significant (p = 0.015) association with AD (Fig. 1). Q-Q plots of the ADSP p-values for the 1,334 variants in CEN_*TREM2*_ and for the remaining 85,383 independent variants with a minor allele count of twenty or more are shown in Fig. 2A demonstrating higher than expected number of significant variants for former.

**Figure 1.**
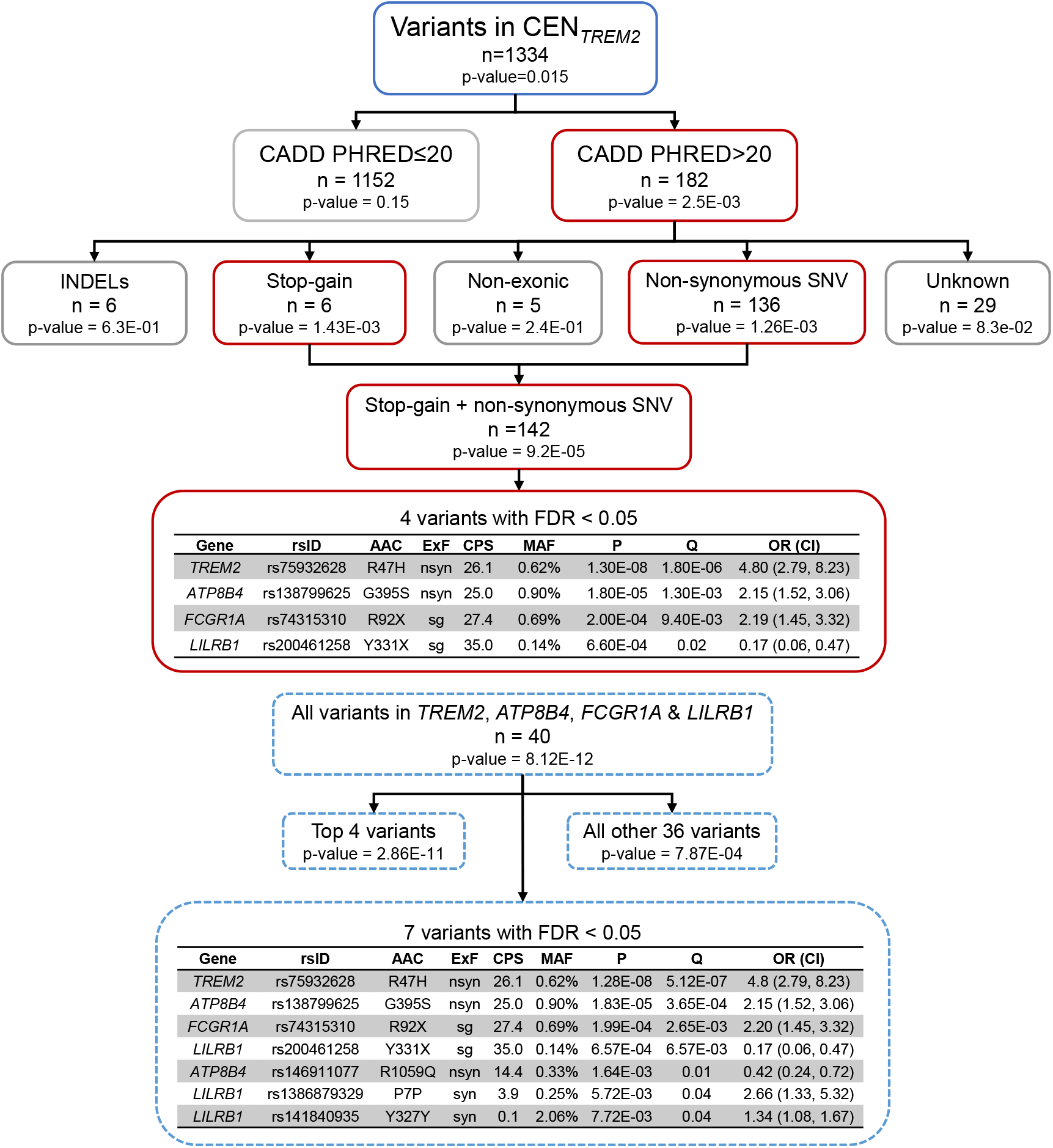
Analytic flow diagram. Variants in the genes of the co-expression network containing *TREM2* (CEN_*TREM2*_) were analyzed in prioritized, sequential steps. PRS analysis was performed using Broad (N=4174) for discovery and the combined WashU plus Baylor (N=5572) as test. The PRS for the 1334 independent variants (r^2^ < 0.2) with a minor allele count of 20 or more in CEN_*TREM*_ showed significant association with AD. The PRS for the 182 deleterious variants in CEN_*TREM2*_ (CADD Phred-scaled scores over 20) showed significant association with AD, whereas the remaining 1152 variants showed only suggestive association. Stratified analysis of the 182 deleterious variants by exonic function showed that association was driven by 136 non-synonymous (nsyn) and 6 stop-gain (sg) variants. False discovery rates (Q) determined after adjustment for 142 deleterious stop-gain and non-synonymous variants showed that *TREM2*.pR47H, *ATP8B4*.pG395S, *FCGR1A*.pR92X, and *LILRB1*.Y331X had significant Q-values. The PRS for these 4 variants showed significant association with AD, as did the 36 remaining variants in *TREM2, ATP8B4, FCGR1A*, and *LILRB1*. The PRS for entire set of 40 variants in the 4 genes showed highly significant, replicable association (Table 3). After adjustment for all 40 variants in *TREM2, ATP8B4, FCGR1A*, and *LILRB1*, which had ADSP p-values ranging from 1.3E-08 to 0.96, 7 variants had significant false discovery rates (Q < 0.05), the four deleterious variants identified above and three additional variants.

**Figure 2.**
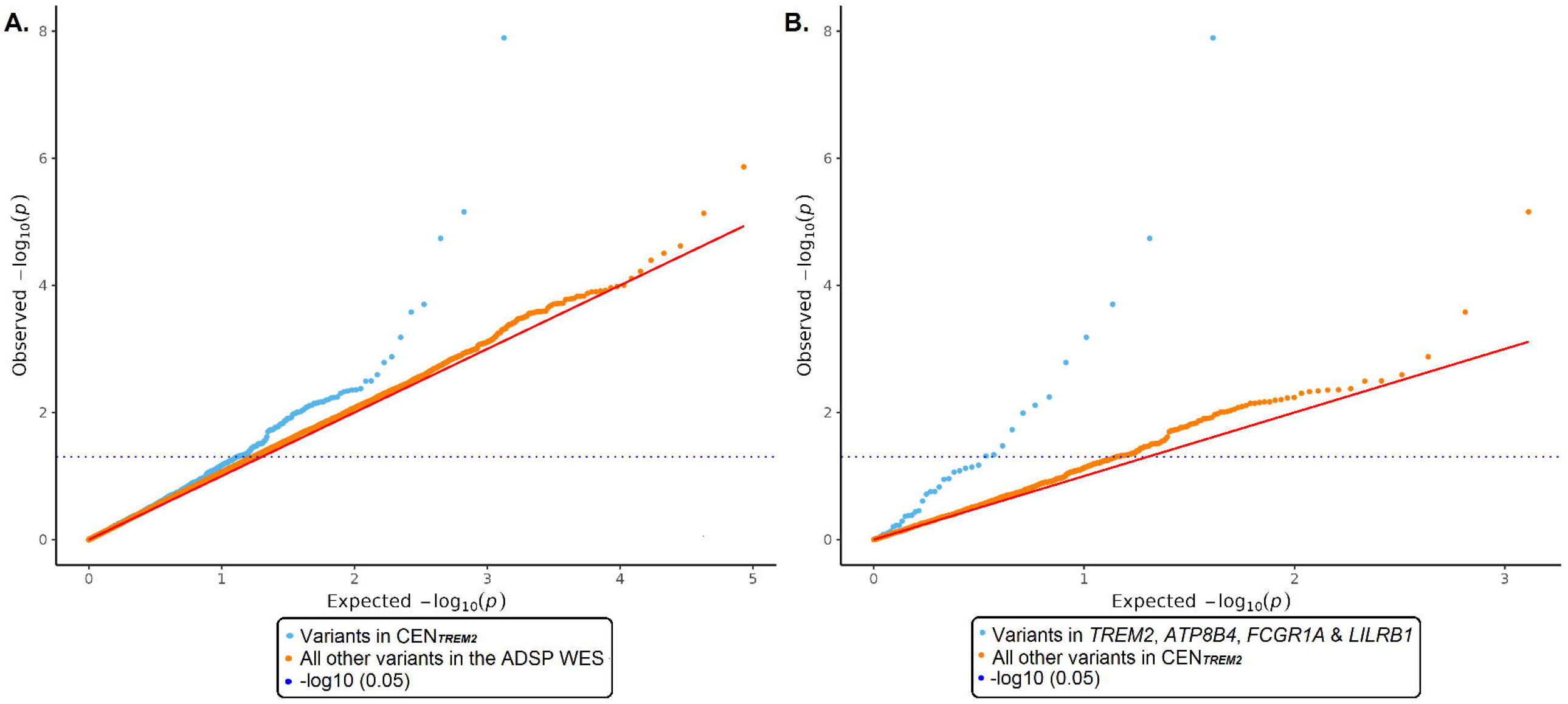
Q-Q plots for ADSP WES variants with minor allele counts (MAC) of 20 or more. Q-Q plots showing p-values from logistic regression adjusting for appropriate covariates for independent variants (r^2^≤0.2) with minor allele count 20 or more in the ADSP (N=9746) dataset. The solid red line shows the p-value distribution expected on the null hypothesis of no association with AD. **A**. ADSP p-values for the 1,334 independent variants located in the 228 genes of CEN_*TREM2*_ (light blue symbols) and the p-values for all remaining independent variants in the ADSP WES dataset (n=85,383; orange symbols) are shown. **B**. ADSP p-values for the 40 variants in the top 4 genes (*TREM2, ATP8B4, FCGR1A, LILRB1*) of CEN_*TREM2*_ (blue symbols) and the p-values for all other variants in CEN_*TREM2*_ (n=1,294; orange symbols) are shown.

### Deleterious CEN_*TREM2*_ variants show significant association with AD

Reasoning that variants judged to be highly deleterious by CADD Phred-scaled scores (CPS) were most likely to have critical impacts on gene function and influence risk of AD, we performed a prioritized analysis of variants in CEN_*TREM2*_ focusing initially on the 182 variants with CPS>20 (Fig. 1). PRS analysis (Table 1) showed that these variants were significantly (p = 2.53×10^−3^) associated with AD, whereas the remaining 1,152 CEN_*TREM2*_ variants showed only suggestive association (p = 0.15). We then analyzed the 182 variants with CPS>20 after stratification by exonic function (Fig. 1, Table 1). Among the 6 subsets, the PRS for stop-gain (p = 1.4×10^−3^) and non-synonymous (p = 1.3×10^−3^) variants showed significant association with AD. The PRS for 142 combined stop-gain and non-synonymous variants further improved the association with AD (p = 9.2×10^−5^). The other subsets showed no evidence of association (Table 1). Note that none of the synonymous variants in CEN_*TREM2*_ had a CPS over 20. To adjust for multiple testing of 5 subsets, false discovery rates (FDR) were determined. Both stop-gain (Q = 3.6×10^−3^) and non-synonymous (Q = 3.6×10^−3^) variants had significant FDR-adjusted p-values (Q-values) for PRS associations.

**Table 1.**
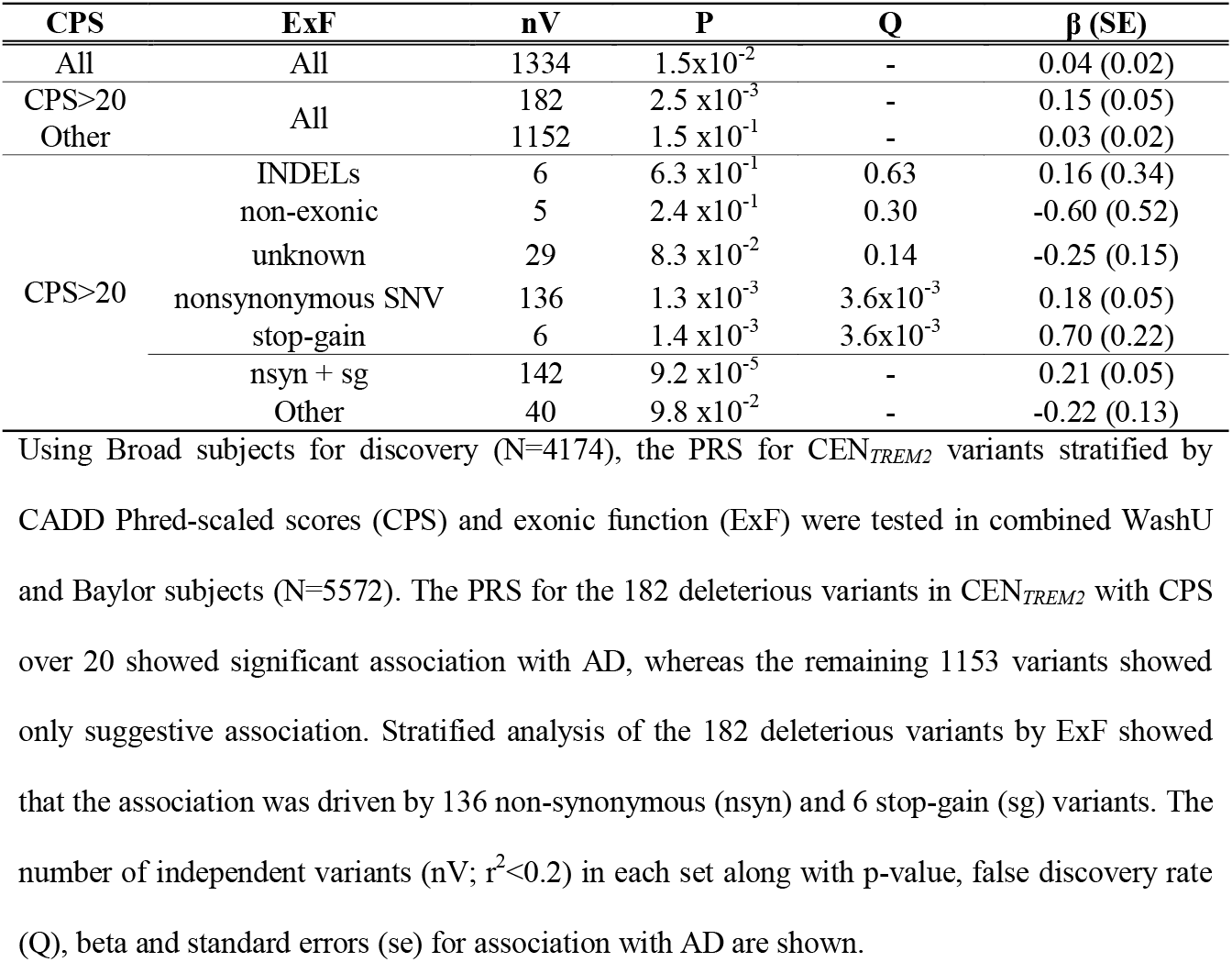
PRS analysis of deleterious variants in CEN_*TREM2*_.

| CPS | ExF | nV | P | Q | $\beta$ (SE) |
| --- | --- | --- | --- | --- | --- |
| All | All | 1334 | $1.5 \times 10^{-2}$ | - | 0.04 (0.02) |
| CPS>20 | All | 182 | $2.5 \times 10^{-3}$ | - | 0.15 (0.05) |
| Other | All | 1152 | $1.5 \times 10^{-1}$ | - | 0.03 (0.02) |
| CPS>20 | INDELs | 6 | $6.3 \times 10^{-1}$ | 0.63 | 0.16 (0.34) |
| | non-exonic | 5 | $2.4 \times 10^{-1}$ | 0.30 | -0.60 (0.52) |
| | unknown | 29 | $8.3 \times 10^{-2}$ | 0.14 | -0.25 (0.15) |
| | nonsynonymous SNV | 136 | $1.3 \times 10^{-3}$ | $3.6 \times 10^{-3}$ | 0.18 (0.05) |
| | stop-gain | 6 | $1.4 \times 10^{-3}$ | $3.6 \times 10^{-3}$ | 0.70 (0.22) |
| | nsyn + sg | 142 | $9.2 \times 10^{-5}$ | - | 0.21 (0.05) |
| | Other | 40 | $9.8 \times 10^{-2}$ | - | -0.22 (0.13) |

Using Broad subjects for discovery (N=4174), the PRS for $CEN_{TREM2}$ variants stratified by CADD Phred-scaled scores (CPS) and exonic function (ExF) were tested in combined WashU and Baylor subjects (N=5572). The PRS for the 182 deleterious variants in $CEN_{TREM2}$ with CPS over 20 showed significant association with AD, whereas the remaining 1153 variants showed only suggestive association. Stratified analysis of the 182 deleterious variants by ExF showed that the association was driven by 136 non-synonymous (nsyn) and 6 stop-gain (sg) variants. The number of independent variants (nV; $r^2 < 0.2$ ) in each set along with p-value, false discovery rate (Q), beta and standard errors (se) for association with AD are shown.

We further restricted the original GWAS to the 142 deleterious, stop-gain and non-synonymous variants identified by our PRS strategy, and using p-values determined from all participants, we calculated the false discovery rate (Q) with respect to this set (Fig. 3). Well annotated results for these 142 variants are shown in Additional File 2. As expected, *TREM2*.pR47H showed highly significant association with AD (Q = 1.8×10^−6^) with an effect size comparable to *APOE* rs429358 (Fig. 3). Deleterious variants in three additional genes (*ATP8B4, FCGR1A*, and *LILRB1*) also showed significant association, including the non-synonymous variant, *ATP8B4*.pG395S (Q = 1.3×10^−3^), and two stop-gain variants, *FCGR1A*.pR92X (Q = 9.4×10^−3^) and *LILRB1*.Y331X (Q = 0.02). Among the 142 deleterious stop-gain and non-synonymous variants, which had p-values ranging from 1.28×10^−8^ – 0.995, there were 11 additional non-synonymous variants with nominally significant P-values (p < 0.05). These variants had false discovery rates ranging from 0.15 to 0.45, and 10 were in genes other than *TREM2, ATP8B4, FCGR1A*, and *LILRB1*. Thus, it is highly likely that additional CEN_*TREM2*_ genes harbor deleterious non-synonymous variants that associate with AD.

**Figure 3.**
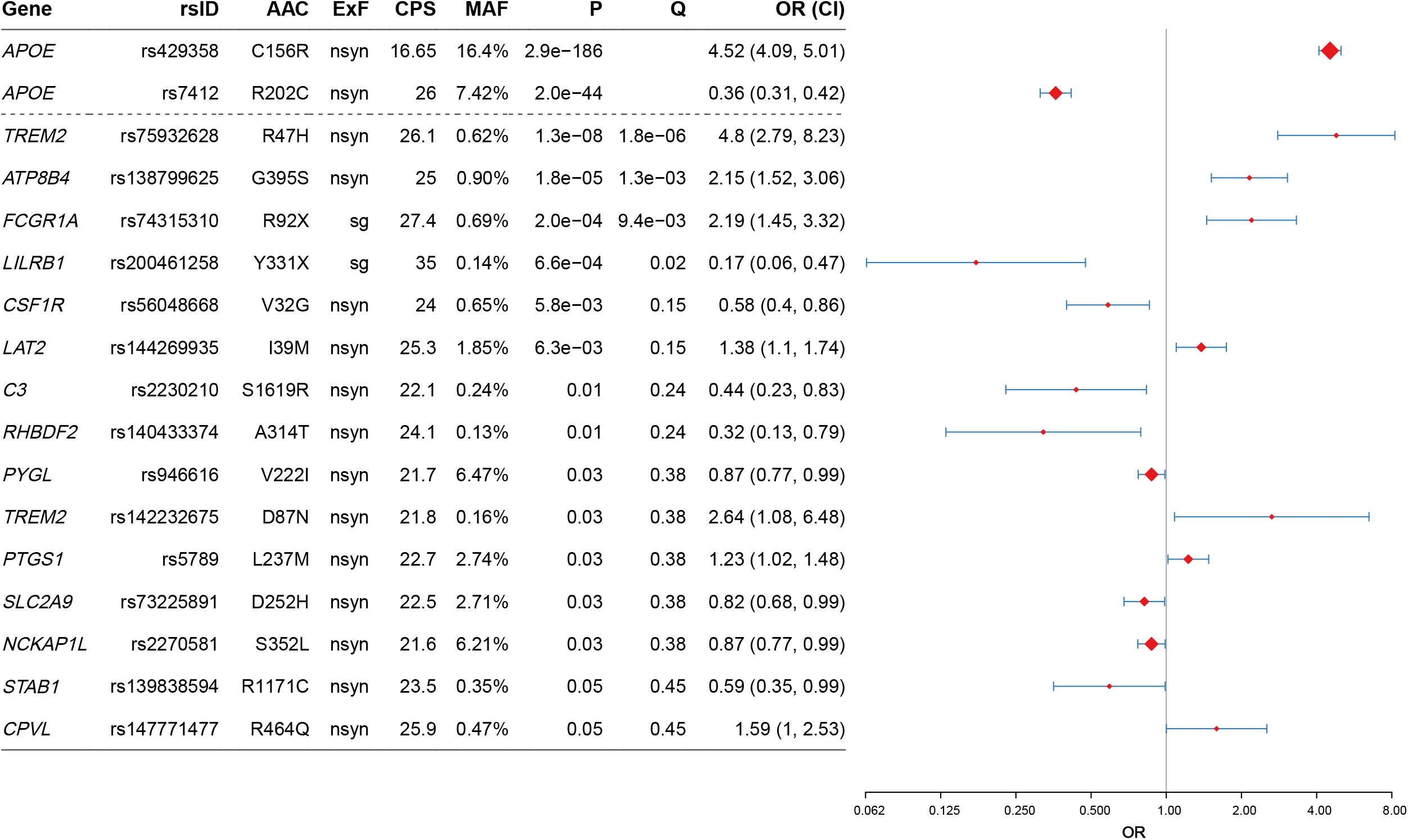
Forest plot of deleterious non-synonymous and stop-gain variants in CEN_*TREM2*_. Annotated results are shown for the 15 variants with ADSP p-values of 0.05 or less. Using all qualifying participants in the ADSP, p-values were obtained for deleterious variants in CEN_*TREM2*_ by logistic regression while adjusting for sex, *APOE* ε2, *APOE* ε4 and the first 3 principal components. After adjustment for the 142 deleterious non-synonymous (nsyn) and stop-gain (sg) variants, which had ADSP p-values ranging from 1.3E-08 to 0.99, 4 variants had significant false discovery rates (Q< 0.05).

### PRS for *TREM2, ATP8B4, FCGR1A*, and *LILRB1* show significant association with AD

Using Broad dataset for discovery, the PRS for the 4 significant, deleterious variants (Fig. 1) showed highly significant association with AD (p = 2.60×10^−11^) in the test dataset comprising combined WashU plus Baylor participants. Remarkably, the PRS for the 36 remaining variants in *TREM2, ATP8B4, FCGR1A*, and *ATP8B4* (Fig. 1) also showed significant association (p=7.87×10^−4^).

When all variants in each gene with a significant deleterious variant were analyzed, the PRS for *TREM2, ATP8B4, FCGR1A*, and *ATP8B4* each showed significant association with AD (Table 2). Q-Q plots of the ADSP p-values for the 40 variants in these 4 genes compared to all other variants in CEN_*TREM2*_ are shown in Fig. 2B demonstrating higher than expected number of observed significant variants.

**Table 2.**
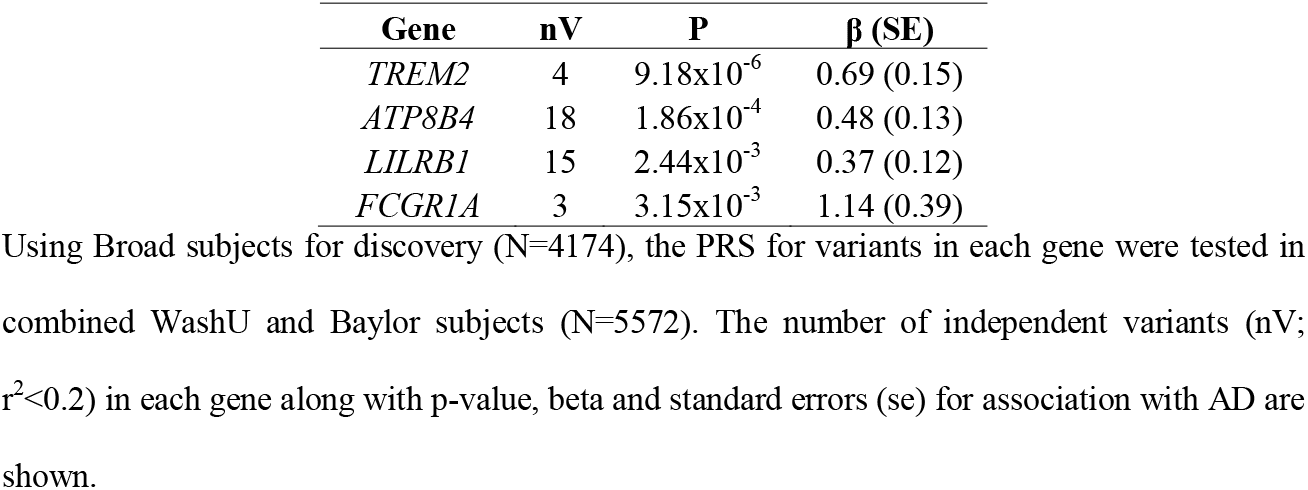
PRS analysis of variants in individual *TREM2, ATP8B4, LILRB1*, and *FCGR1A* genes.

| Gene | nV | P | $\beta$ (SE) |
| --- | --- | --- | --- |
| *TREM2* | 4 | $9.18 \times 10^{-6}$ | 0.69 (0.15) |
| *ATP8B4* | 18 | $1.86 \times 10^{-4}$ | 0.48 (0.13) |
| *LILRB1* | 15 | $2.44 \times 10^{-3}$ | 0.37 (0.12) |
| *FCGR1A* | 3 | $3.15 \times 10^{-3}$ | 1.14 (0.39) |

Using Broad subjects for discovery (N=4174), the PRS for variants in each gene were tested in combined WashU and Baylor subjects (N=5572). The number of independent variants (nV; $r^2 < 0.2$ ) in each gene along with p-value, beta and standard errors (se) for association with AD are shown.

When tested in the WashU and Baylor datasets individually, as well as in the combined datasets, the PRS for all 40 variants in the four genes (Table 3) showed significant association in WashU [p = 5.40×10^−8^, β (SE) = 0.53 (0.10)], which replicated in Baylor [p = 6.87×10^−5^, β (SE) = 0.48 (0.12)], and became highly significant in the combined dataset [p = 8.12×10^−12^, β (SE) = 0.52 (0.08)]. We then analyzed the receiver operator characteristics (ROC) curve to determine the improvement in AUC (Δ_AUC_) for models that included PRS, sex, *APOE* ε2, *APOE* ε4 and the first three principal components as compared to models without PRS. This ROC analysis showed that the PRS for all 40 variants improved the area under the curve in the combined WashU plus Baylor test dataset by 0.75% (Table 3). After adjustment for all 40 variants in *TREM2, ATP8B4, FCGR1A*, and *LILRB1*, which had ADSP p-values ranging from 1.28×10^−8^ to 0.961, 7 variants had significant false discovery rates (Q < 0.05), the four deleterious variants identified above and three additional variants (Fig. 1, 4). Well annotated results for these 40 variants are shown in Additional File 3.

**Table 3.** PRS analysis of combined variants from *TREM2, ATP8B4, LILRB1*, and *FCGR1A* genes.

| Genes | nV | Test Set | P | $\beta$ (SE) | $\Delta_{AUC}$ |
| --- | --- | --- | --- | --- | --- |
| *TREM2*, *ATP8B4*, *LILRB1*,<br>*FCGR1A* | 40 | WashU | $5.40 \times 10^{-8}$ | 0.53 (0.10) | 0.84% |
| | | Baylor | $6.87 \times 10^{-5}$ | 0.48 (0.12) | 0.71% |
| | | Washu+Baylor | $8.12 \times 10^{-12}$ | 0.52 (0.08) | 0.75% |
| *ATP8B4*, *LILRB1*, *FCGR1A* | 36 | WashU | $1.45 \times 10^{-5}$ | 0.48 (0.11) | 0.68% |
| | | Baylor | $2.35 \times 10^{-3}$ | 0.43 (0.14) | 0.58% |
| | | Washu+Baylor | $9.89 \times 10^{-8}$ | 0.46 (0.09) | 0.59% |

Using Broad subjects for discovery (N=4174), the PRS for the combined set of 40 variants from *TREM2*, *ATP8B4*, *LILRB1*, and *FCGR1A* or the set of 36 variants from the 3 novel AD genes (*ATP8B4*, *LILRB1*, and *FCGR1A*) was tested in WashU (N=3299), Baylor (N=2273), and combined WashU plus Baylor subjects (N=5572). The number of independent variants (nV; $r^2 < 0.2$ ) in each set along with p-value, beta, standard errors (se). Improvement in AUC ( $\Delta_{AUC}$ ) is shown for the model that included PRS, sex, *APOE* $\epsilon 2$ , *APOE* $\epsilon 4$ and the first three principal components as compared to the same model without PRS.

**Figure 4.**
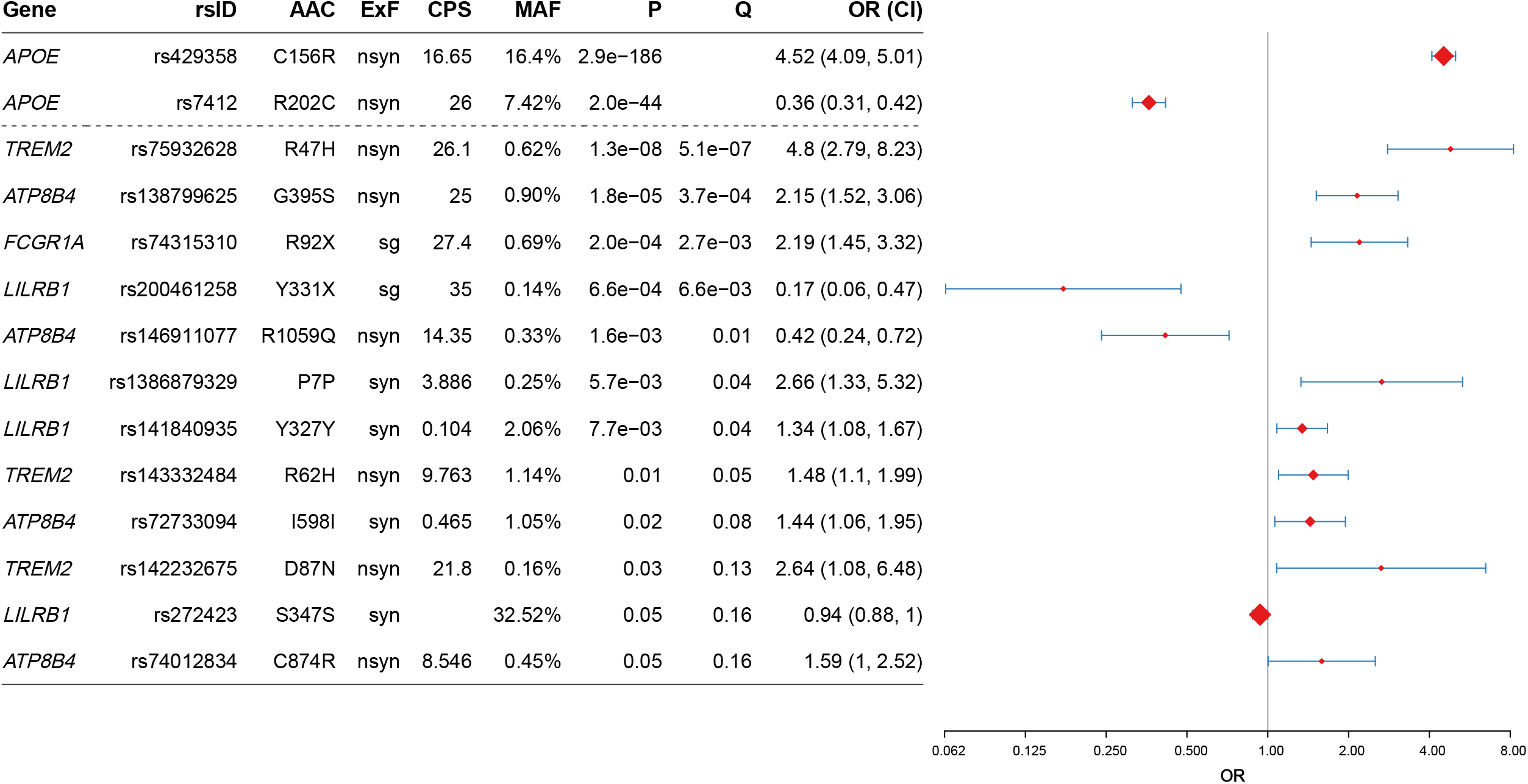
Forest plot of variants from *TREM2, ATP8B4, LILRB1*, and *FCGR1A* genes. Annotated results are shown for the 12 variants in *TREM2, ATP8B4, FCGR1A*, and *LILRB1* with ADSP P-values of 0.05 or less. Using all qualifying participants in the ADSP, p-values were obtained for all variants in the four genes by logistic regression while adjusting for sex, *APOE* ε2, *APOE* ε4 and the first 3 principal components. After adjustment for all 40 variants in these 4 genes, which had ADSP P-values ranging from 1.3E-08 to 0.96, 7 variants had significant false discovery rates (Q< 0.05).

Importantly, the PRS for all 36 variants in the 3 novel genes, (*ATP8B4, FCGR1A*, and *LILRB1*; Table 3) also showed significant association in the WashU dataset [p = 1.45×10^−5^, β (SE) = 0.48 (0.11)], which replicated in Baylor [p = 2.53×10^−3^, β (SE) = 0.43 (0.14)] and became highly significant in the combined dataset [p = 9.89×10^−8^, β (SE) = 0.46 (0.09)]. The PRS for these 36 variants accounted for 0.59% of the area under the ROC curve (Table 3) in the combined WashU plus Baylor test participants. These findings demonstrate that CEN_*TREM2*_ harbors AD risk variants in not only *TREM2* but also three novel immune pathway genes.

## Discussion

In this study, we show that the PRS for the 142 deleterious (CPS>20) non-synonymous and stop-gain variants in CEN_*TREM2*_ is significantly associated with AD (p = 9.20×10^−5^) and that 4 variants in this polygenic component are also individually significant after FDR adjustment. Each of these variants is a deleterious, protein-altering variant with a MAF of 0.1 - 1.0%. All four variants reside in genes that, in the brain, are selectively expressed in microglial cells [49, 50], and have strong effects on risk of AD comparable to those of the well-known *APOE* alleles (Fig. 4). The strong association of *TREM2*.pR47H with AD is well-established [9-16]. One study, posted on medRxiv, assessing gene-based burden of rare damaging variants in exome sequencing data has reported that exonic variants in *ATP8B4* associate with AD, an association that was driven mainly by the same *ATP8B4*.pG395S variant described here [51]. The stop-gain variants *FCGR1A*.pR92X and *LILRB1*pY331X have not previously been reported to associate with AD.

The PRS for all 40 variants in *TREM2, ATP8B4, FCGR1A*, and *LILRB1* showed significant, replicable association with AD (Table 3), as did the PRS for all 36 variants in *ATP8B4, FCGR1A*, and *LILRB1*, the 3 novel genes identified here. After adjustment for the 40 variants in *TREM2, ATP8B4, FCGR1A*, and *LILRB1*, 7 variants had significant association after adjusting for multiple testing (FDR) – the four identified above, one additional variant in *ATP8B4*, and two additional variants in *LILRB1* (Fig. 1, 4). The additional *ATP8B4* variant is a non-synonymous variant, *ATP8B4*.pR1059Q, with a CPS of 14.4 and an OR of 0.42 (0.24, 0.72) showing association with *decreased* risk of AD with an effect size comparable to that of the protective *APOE* ε2 allele (Fig. 4). The additional *LILRB1* variants are synonymous variants, which are associated with increased risk of AD. Many genes have variants in intronic and flanking regions that alter gene expression or splicing. Among the many synonymous exonic variants, some will be linked to flanking or intronic variants with functional effects. Thus, the synonymous variants in *LILRB1* may well show significant association with increased risk of AD through linkage to nearby functional variants.

*ATP8B4* encodes a protein that is a component of a P4-ATPase flippase complex, which catalyzes the hydrolysis of ATP coupled to the transport of aminophospholipids from the outer to the inner leaflet of various membranes and ensures the maintenance of asymmetric distribution of phospholipids [52]. Phospholipid translocation seems also to be implicated in vesicle formation and in uptake of lipid signaling molecules [53].

The stop-gain variants in *FCGR1A* and *LILRB1* will almost certainly cause a loss of function. *FCGR1A* is a high affinity receptor for the Fc region of immunoglobulin gamma that functions in both innate and adaptive immunity [54]. Just as signaling downstream of *TREM2* is mediated by *TYROBP* which contains an immunoreceptor tyrosine-based activation motif (ITAM), ligand binding to *FCGR1A* signals through the ITAM containing *FCGER* protein. This suggests that the biological action of *TREM2* variants and a stop gain variant in *FCGR1A* may be very similar. A previous study identified *FCGR1A*.pR92X in an apparently healthy family of individuals who lacked phagocyte expression of the IgG receptor (CD64) and were unable to support mouse anti-CD3-induced induced T cell mitogenesis. The premature introduction of a stop codon was associated with a decrease in abundance of FcyRIa mRNA in individuals homozygous for the mutation [55]. Thus, it appears that appropriate activation of *FCGR1A* can reduce risk of AD. *LILRB1* belongs to the subfamily B class of LIR receptors. It is expressed on immune cells where it is known to bind to MHC class I molecules on antigen presenting cells and transduces a negative signal though immunoreceptor tyrosine-based inhibition motif (ITIM) that suppresses immune responses [56, 57]. *LILRB1* on natural killer cells may also have a prominent role in inflammation following an immune response [58]. The stop-gain variant in *LILRB1*, which strongly decreases risk of AD, will likely impair the effect of ligands that modulate the immune system by binding to *LILRB1*. Thus, it appears that activation of *LILRB1* can increase risk of AD. An important implication of this is that well-designed, appropriately targeted *LILRB1* inhibitors might have a strong beneficial effect on AD pathogenesis.

In AD, the concept that neuroinflammation was harmful and that proinflammatory stimuli would increase risk for AD dominated discussion of the role of the innate immune systems in AD for many years and served as the rationale for several clinical trials [59]. However, this concept has been challenged by both genetic and modeling data as well as results from failed clinical studies of anti-inflammatory agents in AD (reviewed in [8]). Notably, the directionality of risk associated with variants in immune genes identified in this study is entirely consistent with risk associated with *TREM2* and *PLCG2* variants [60-63]. Indeed, our current data and previous data suggest that increased activity (or increased expression) of immune activating proteins will decrease risk for AD as will loss of function (or decreased expression) of immune suppressing proteins, and conversely that increased activity (or increased expression) of immune inhibiting proteins will confer increased risk for AD as will loss of function (or decreased expression) of immune activating proteins. Of course, biology rarely works in completely dichotomous fashion and immune signaling pathways are often tightly regulated with the overall impact of a variant on a given signaling pathway likely to be contextually dependent. Thus, experimental data will be needed to understand how these newly identified variants impact AD-relevant phenotypes and innate immune signaling pathways.

Virtually all genes encode proteins that are pleiotropic. For this reason, virtually every deleterious variant that associates with AD will have multiple effects, only some of which play an important role in altering risk of AD. In this study, we identify a set of variants in *TREM2* and in 3 novel immune pathway genes with powerful effects that both increase and decrease risk of AD. Concerted studies of these powerful variants should be pursued, as they have the potential to identify novel therapeutic targets within the immune system in the same way that study of *APOE* variants and the *APP, PSEN1*, and *PSEN2* mutations that cause early onset familial AD identified Aβ, more specifically Aβ42 as a therapeutic target for AD.

The work described here was undertaken as part of a multi-site study aimed at identifying novel therapeutic targets for AD. Because the innate immune system has an important role in AD, our role was to search in innate immune genes for novel genetic variants associated with AD. When we analyzed the deleterious variants in all 19 co-expression modules identified by Allen et al [46], PRS for CEN_*TREM2*_ was the most significantly associated with AD risk (p =2.5×10^−3^, Q = 0.034). Thus, there was a strong functional and statistical rationale for focusing on deleterious variants in CEN_*TREM2*_ genes. This focus enabled us to obtain strong evidence that multiple variants in *TREM2, ATP8B4, FCGR1A*, and *LILRB1* show strong association with AD. It is nevertheless important that follow up studies be performed both to confirm the results reported here and to narrow the 95% CIs thereby better defining effect size.

## Conclusions

Our study demonstrates the practical utility of combining information on the genes in a co-expression network with genetic risk association data in order to perform a functionally focused analyses of deleterious variants in the genes of a candidate disease pathway. Using this approach, we identify three novel immune pathway genes (*ATP8B4, LILRB1*, and *FCGR1A*) with a variant that shows highly significant (P < 0.001, Q < 0.01) association with AD. Like *TREM2*.pR47H, each of these variants has a minor allele frequency less than 1% and is a deleterious, protein altering variant with a strong effect that increases or decreases (*LILRB1*.pY331X) risk of AD. Additional variants in these genes also alter risk of AD. The variants identified here are ideally suited for studies aimed at understanding how the immune system may be modulated to alter risk of AD.

## Data Availability

All data produced in the present study are available upon reasonable request to the authors

https://www.niagads.org/adsp/content/home

## List of abbreviations

Aβ: amyloid β protein
AD: Alzheimer’s disease
ADSP: Alzheimer’s Disease Sequencing Project
*ATP8B4*: gene encoding probable phospholipid-transporting ATPase IM
AUC: area-under-the-curve
CADD: Combined Annotation Dependent Depletion
CEN_TREM2_: co-expression network containing TREM2
CPS: Combined Annotation Dependent Depletion Phred-scaled score
*FCGR1A*: gene encoding Fc gamma receptor 1A
*LILRB1*: gene encoding leukocyte immunoglobulin-like receptor subfamily B member 1
MAC: minor allele count
MAF: minor allele frequency
PC: principal component
PRS: polygenic risk score
QC: quality control
ROC: receiver operator characteristics
SE: standard error
*TREM2*: gene encoding triggering receptor expressed on myeloid cells 2
WES: whole exome sequence
WGCNA: weighted gene co-expression network analysis.

## Declarations

### Ethics approval and consent to participate

Participants or their caregivers provided written informed consent in the original studies. The current study protocol was granted an exemption by the Mayo Clinic Institutional Review Board because the analyses were carried out on de-identified, off-the-shelf data; therefore, additional informed consent was not required.

### Consent for publication

Not applicable

### Availability of data and materials

Whole exome sequencing (WES) data and corresponding clinical information utilized in this study can be obtained from the National Institute of Aging Genetics of Alzheimer’s Disease Data Storage Service here: https://www.niagads.org/adsp/content/home.The R code used during the current study is available from the corresponding author on reasonable request.

## Authors’ contributions

JSR and SGY conceived the project and performed all the analyses. NET, XW, MA and MMC contributed RNAseq data and WGCNA analyses. BJC, JMB and GDJ provided input on QC and PRS analyses. JSR, SGY and TEG prepared the first draft of the manuscript. OB assisted with subsequent drafts. All authors contributed to the final manuscript.

## Acknowledgements

We thank the patients and their families for their participation, without which these studies would not have been possible.

For samples collected through the Sun Health Research Institute Brain and Body Donation Program of Sun City, Arizona and utilized in the brain expression studies: The Brain and Body Donation Program is supported by the National Institute of Neurological Disorders and Stroke (U24NS072026 National Brain and Tissue Resource for Parkinson’s Disease and Related Disorders), the National Institute on Aging (P30 AG19610 Arizona Alzheimer’s Disease Core Center), the Arizona Department of Health Services (contract 211002, Arizona Alzheimer’s Research Center), the Arizona Biomedical Research Commission (contracts 4001, 0011, 05-901, and 1001 to the Arizona Parkinson’s Disease Consortium), and the Michael J. Fox Foundation for Parkinson’s Research.

## Acknowledgment Statement for the ADSP

The Alzheimer’s Disease Sequencing Project (ADSP) is comprised of two Alzheimer’s Disease (AD) genetics consortia and three National Human Genome Research Institute (NHGRI) funded Large Scale Sequencing and Analysis Centers (LSAC). The two AD genetics consortia are the Alzheimer’s Disease Genetics Consortium (ADGC) funded by NIA (U01 AG032984), and the Cohorts for Heart and Aging Research in Genomic Epidemiology (CHARGE) funded by NIA (R01 AG033193), the National Heart, Lung, and Blood Institute (NHLBI), other National Institute of Health (NIH) institutes and other foreign governmental and non-governmental organizations. The Discovery Phase analysis of sequence data is supported through UF1AG047133 (to Drs. Schellenberg, Farrer, Pericak-Vance, Mayeux, and Haines); U01AG049505 to Dr. Seshadri; U01AG049506 to Dr. Boerwinkle; U01AG049507 to Dr. Wijsman; and U01AG049508 to Dr. Goate and the Discovery Extension Phase analysis is supported through U01AG052411 to Dr. Goate, U01AG052410 to Dr. Pericak-Vance and U01 AG052409 to Drs. Seshadri and Fornage.

Sequencing for the Follow Up Study (FUS) is supported through U01AG057659 (to Drs. PericakVance, Mayeux, and Vardarajan) and U01AG062943 (to Drs. Pericak-Vance and Mayeux). Data generation and harmonization in the Follow-up Phase is supported by U54AG052427 (to Drs. Schellenberg and Wang). The FUS Phase analysis of sequence data is supported through U01AG058589 (to Drs. Destefano, Boerwinkle, De Jager, Fornage, Seshadri, and Wijsman), U01AG058654 (to Drs. Haines, Bush, Farrer, Martin, and Pericak-Vance), U01AG058635 (to Dr. Goate), RF1AG058066 (to Drs. Haines, Pericak-Vance, and Scott), RF1AG057519 (to Drs. Farrer and Jun), R01AG048927 (to Dr. Farrer), and RF1AG054074 (to Drs. Pericak-Vance and Beecham).

The ADGC cohorts include: Adult Changes in Thought (ACT) (UO1 AG006781, UO1 HG004610, UO1 HG006375, U01 HG008657), the Alzheimer’s Disease Centers (ADC) (P30 AG019610, P30 AG013846, P50 AG008702, P50 AG025688, P50 AG047266, P30 AG010133, P50 AG005146, P50 AG005134, P50 AG016574, P50 AG005138, P30 AG008051, P30 AG013854, P30 AG008017, P30 AG010161, P50 AG047366, P30 AG010129, P50 AG016573, P50 AG016570, P50 AG005131, P50 AG023501, P30 AG035982, P30 AG028383, P30 AG010124, P50 AG005133, P50 AG005142, P30 AG012300, P50 AG005136, P50 AG033514, P50 AG005681, and P50 AG047270), the Chicago Health and Aging Project (CHAP) (R01 AG11101, RC4 AG039085, K23 AG030944), Indianapolis Ibadan (R01 AG009956, P30 AG010133), the Memory and Aging Project (MAP) (R01 AG17917), Mayo Clinic (MAYO) (R01 AG032990, U01 AG046139, R01 NS080820, RF1 AG051504, P50 AG016574), Mayo Parkinson’s Disease controls (NS039764, NS071674, 5RC2HG005605), University of Miami (R01 AG027944, R01 AG028786, R01 AG019085, IIRG09133827, A2011048), the Multi-Institutional Research in Alzheimer’s Genetic Epidemiology Study (MIRAGE) (R01 AG09029, R01 AG025259), the National Cell Repository for Alzheimer’s Disease (NCRAD) (U24 AG21886), the National Institute on Aging Late Onset Alzheimer’s Disease Family Study (NIA-LOAD) (R01 AG041797), the Religious Orders Study (ROS) (P30 AG10161, R01 AG15819), the Texas Alzheimer’s Research and Care Consortium (TARCC) (funded by the Darrell K Royal Texas Alzheimer’s Initiative), Vanderbilt University/Case Western Reserve University (VAN/CWRU) (R01 AG019757, R01 AG021547, R01 AG027944, R01 AG028786, P01 NS026630, and Alzheimer’s Association), the Washington Heights-Inwood Columbia Aging Project (WHICAP) (RF1 AG054023), the University of Washington Families (VA Research Merit Grant, NIA: P50AG005136, R01AG041797, NINDS: R01NS069719), the Columbia University HispanicEstudio Familiar de Influencia Genetica de Alzheimer (EFIGA) (RF1 AG015473), the University of Toronto (UT) (funded by Wellcome Trust, Medical Research Council, Canadian Institutes of Health Research), and Genetic Differences (GD) (R01 AG007584). The CHARGE cohorts are supported in part by National Heart, Lung, and Blood Institute (NHLBI) infrastructure grant HL105756 (Psaty), RC2HL102419 (Boerwinkle) and the neurology working group is supported by the National Institute on Aging (NIA) R01 grant AG033193.

The CHARGE cohorts participating in the ADSP include the following: Austrian Stroke Prevention Study (ASPS), ASPS-Family study, and the Prospective Dementia Registry-Austria (ASPS/PRODEM-Aus), the Atherosclerosis Risk in Communities (ARIC) Study, the Cardiovascular Health Study (CHS), the Erasmus Rucphen Family Study (ERF), the Framingham Heart Study (FHS), and the Rotterdam Study (RS). ASPS is funded by the Austrian Science Fond (FWF) grant number P20545-P05 and P13180 and the Medical University of Graz. The ASPS-Fam is funded by the Austrian Science Fund (FWF) project I904),the EU Joint Programme - Neurodegenerative Disease Research (JPND) in frame of the BRIDGET project (Austria, Ministry of Science) and the Medical University of Graz and the Steiermärkische Krankenanstalten Gesellschaft. PRODEM-Austria is supported by the Austrian Research Promotion agency (FFG) (Project No. 827462) and by the Austrian National Bank (Anniversary Fund, project 15435. ARIC research is carried out as a collaborative study supported by NHLBI contracts (HHSN268201100005C, HHSN268201100006C, HHSN268201100007C, HHSN268201100008C, HHSN268201100009C, HHSN268201100010C, HHSN268201100011C, and HHSN268201100012C). Neurocognitive data in ARIC is collected by U01 2U01HL096812, 2U01HL096814, 2U01HL096899, 2U01HL096902, 2U01HL096917 from the NIH (NHLBI, NINDS, NIA and NIDCD), and with previous brain MRI examinations funded by R01-HL70825 from the NHLBI. CHS research was supported by contracts HHSN268201200036C, HHSN268200800007C, N01HC55222, N01HC85079, N01HC85080, N01HC85081, N01HC85082, N01HC85083, N01HC85086, and grants U01HL080295 and U01HL130114 from the NHLBI with additional contribution from the National Institute of Neurological Disorders and Stroke (NINDS). Additional support was provided by R01AG023629, R01AG15928, and R01AG20098 from the NIA. FHS research is supported by NHLBI contracts N01-HC-25195 and HHSN268201500001I. This study was also supported by additional grants from the NIA (R01s AG054076, AG049607 and AG033040 and NINDS (R01 NS017950). The ERF study as a part of EUROSPAN (European Special Populations Research Network) was supported by European Commission FP6 STRP grant number 018947 (LSHG-CT-2006-01947) and also received funding from the European Community’s Seventh Framework Programme (FP7/2007-2013)/grant agreement HEALTH-F4-2007-201413 by the European Commission under the programme “Quality of Life and Management of the Living Resources” of 5th Framework Programme (no. QLG2-CT-2002-01254). High-throughput analysis of the ERF data was supported by a joint grant from the Netherlands Organization for Scientific Research and the Russian Foundation for Basic Research (NWO-RFBR 047.017.043). The Rotterdam Study is funded by Erasmus Medical Center and Erasmus University, Rotterdam, the Netherlands Organization for Health Research and Development (ZonMw), the Research Institute for Diseases in the Elderly (RIDE), the Ministry of Education, Culture and Science, the Ministry for Health, Welfare and Sports, the European Commission (DG XII), and the municipality of Rotterdam. Genetic data sets are also supported by the Netherlands Organization of Scientific Research NWO Investments (175.010.2005.011, 911-03-012), the Genetic Laboratory of the Department of Internal Medicine, Erasmus MC, the Research Institute for Diseases in the Elderly (014-93-015; RIDE2), and the Netherlands Genomics Initiative (NGI)/Netherlands Organization for Scientific Research (NWO) Netherlands Consortium for Healthy Aging (NCHA), project 050-060-810. All studies are grateful to their participants, faculty and staff. The content of these manuscripts is solely the responsibility of the authors and does not necessarily represent the official views of the National Institutes of Health or the U.S. Department of Health and Human Services.

The FUS cohorts include: the Alzheimer’s Disease Centers (ADC) (P30 AG019610, P30 AG013846, P50 AG008702, P50 AG025688, P50 AG047266, P30 AG010133, P50 AG005146, P50 AG005134, P50 AG016574, P50 AG005138, P30 AG008051, P30 AG013854, P30 AG008017, P30 AG010161, P50 AG047366, P30 AG010129, P50 AG016573, P50 AG016570, P50 AG005131, P50 AG023501, P30 AG035982, P30 AG028383, P30 AG010124, P50 AG005133, P50 AG005142, P30 AG012300, P50 AG005136, P50 AG033514, P50 AG005681, and P50 AG047270), Alzheimer’s Disease Neuroimaging Initiative (ADNI) (U19AG024904), Amish Protective Variant Study (RF1AG058066), Cache County Study (R01AG11380, R01AG031272, R01AG21136, RF1AG054052), Case Western Reserve University Brain Bank (CWRUBB) (P50AG008012), Case Western Reserve University Rapid Decline (CWRURD) (RF1AG058267, NU38CK000480), CubanAmerican Alzheimer’s Disease Initiative (CuAADI) (3U01AG052410), Estudio Familiar de Influencia Genetica en Alzheimer (EFIGA) (5R37AG015473, RF1AG015473, R56AG051876), Genetic and Environmental Risk Factors for Alzheimer Disease Among African Americans Study (GenerAAtions) (2R01AG09029, R01AG025259, 2R01AG048927), Gwangju Alzheimer and Related Dementias Study (GARD) (U01AG062602), Hussman Institute for Human Genomics Brain Bank (HIHGBB) (R01AG027944, Alzheimer’s Association “Identification of Rare Variants in Alzheimer Disease”), Ibadan Study of Aging (IBADAN) (5R01AG009956), Mexican Health and Aging Study (MHAS) (R01AG018016), Multi-Institutional Research in Alzheimer’s Genetic Epidemiology (MIRAGE) (2R01AG09029, R01AG025259, 2R01AG048927), Northern Manhattan Study (NOMAS) (R01NS29993), Peru Alzheimer’s Disease Initiative (PeADI) (RF1AG054074), Puerto Rican 1066 (PR1066) (Wellcome Trust (GR066133/GR080002), European Research Council (340755)), Puerto Rican Alzheimer Disease Initiative (PRADI) (RF1AG054074), Reasons for Geographic and Racial Differences in Stroke (REGARDS) (U01NS041588), Research in African American Alzheimer Disease Initiative (REAAADI) (U01AG052410), Rush Alzheimer’s Disease Center (ROSMAP) (P30AG10161, R01AG15819, R01AG17919), University of Miami Brain Endowment Bank (MBB), and University of Miami/Case Western/North Carolina A&T African American (UM/CASE/NCAT) (U01AG052410, R01AG028786).

The four LSACs are: the Human Genome Sequencing Center at the Baylor College of Medicine (U54 HG003273), the Broad Institute Genome Center (U54HG003067), The American Genome Center at the Uniformed Services University of the Health Sciences (U01AG057659), and the Washington University Genome Institute (U54HG003079).

Biological samples and associated phenotypic data used in primary data analyses were stored at Study Investigators institutions, and at the National Cell Repository for Alzheimer’s Disease (NCRAD, U24AG021886) at Indiana University funded by NIA. Associated Phenotypic Data used in primary and secondary data analyses were provided by Study Investigators, the NIA funded Alzheimer’s Disease Centers (ADCs), and the National Alzheimer’s Coordinating Center (NACC, U01AG016976) and the National Institute on Aging Genetics of Alzheimer’s Disease Data Storage Site (NIAGADS, U24AG041689) at the University of Pennsylvania, funded by NIA This research was supported in part by the Intramural Research Program of the National Institutes of health, National Library of Medicine. Contributors to the Genetic Analysis Data included Study Investigators on projects that were individually funded by NIA, and other NIH institutes, and by private U.S. organizations, or foreign governmental or nongovernmental organizations.

## Additional files

**Additional file 1. Sample demographics by AD cases and cognitively normal controls** Total number of participants, number and percentage of females, age and standard deviation (sd), and the number and percentage of *APOE* ε4 positives is shown for each subset. File Format: .xlsx

**Additional File 2. Annotated results are shown for the 142 deleterious non-synonymous and stop-gain variants in CEN**_***TREM2***_ Using all qualifying participants in the ADSP, p-values were obtained for deleterious variants in CEN_*TREM2*_ by logistic regression while adjusting for sex, *APOE* ε2, *APOE* ε4 and the first 3 principal components. After adjustment for the 142 variants in this polygenic component, which had ADSP p-values ranging from 1.28×10^−8^ to 0.995, 4 variants had significant false discovery rates (Q< 0.05). File Format: .xlsx

**Additional File 3. Annotated results for all independent variants (r**^**2**^**<0.2) in *TREM2, ATP8B4, FCGR1A*, and *LILRB1*** Using all qualifying participants in the ADSP, p-values were obtained for all variants in the four genes by logistic regression while adjusting for sex, *APOE* ε2, *APOE* ε4 and the first 3 principal components. After adjustment for all 40 variants in these 4 genes, which had ADSP p-values ranging from 1.3×10^−8^ to 0.96, 7 variants had significant false discovery rates (Q< 0.05). File Format: .xlsx

